# Equity, cost and disability adjusted life years of tuberculosis treatment supported by digital adherence technologies and differentiated care in Ethiopia: a trial-based distributional cost-effectiveness analysis

**DOI:** 10.1101/2024.07.28.24310767

**Authors:** Nicola Foster, Amare W Tadesse, Mahilet Belachew, Mamush Sahlie, Christopher Finn McQuaid, Lara Goscé, Ahmed Bedru, Tofik Abdurhman, Demekech G Umeta, Amanuel Shiferaw, Gedion T Weldemichael, Taye Letta Janfa, Norma Madden, Salome Charalambous, Job van Rest, Kristian van Kalmthout, Degu Jerene, Katherine L Fielding

## Abstract

**Summary:** *Background:* Evidence of the cost-effectiveness of digital adherence technologies (DATs) for supporting tuberculosis treatment has been inconclusive and primarily omitted patient-incurred costs. We aimed to assess the societal costs, equity impact and cost-effectiveness of DATs and differentiated care compared to routine care in Ethiopia.

*Methods:* We conducted a distributional cost-effectiveness analysis using data from the cluster randomised trial that evaluated the implementation of labels and pillbox followed by differentiated care to support tuberculosis treatment adherence in 78 health facilities in Ethiopia. We estimated the costs, cost per disability-adjusted life year (DALYs) averted and equity impact of the implementation of the DATs interventions. Costs and DALYs were estimated at a participant level based on patient events collected during the trial and the trial endpoints for intention-to-treat population. Uncertainty in cost-effectiveness estimates were assessed by plotting cost-effectiveness acceptability frontiers. The trial is registered with Pan African Clinical Trial Registry (PACTR) PACTR202008776694999, registered on 11 August 2020 at https://pactr.samrc.ac.za/TrialDisplay.aspx?TrialID=12241 and has been completed.

*Findings:* The mean total societal treatment cost per trial participant was US$507 (95%CI: 458; 555) in the SOC, US$196 (95%CI: 190; 218) in the labels and US$206 (95%CI: 167; 213) in the pillbox study arms. We estimated that there was a 49-56% probability that the implementation of the DAT interventions, would improve the cost-effectiveness of tuberculosis treatment at a cost-effectiveness threshold of US$100. There was no difference in DALYs between socio-economic position groups (*p*=0.920), however, patient costs were less concentrated among those relatively poor in the intervention arms – labels (illness concentration index [ICI]=0.03 (95%CI: 0.01; 0.05)) and pillbox (ICI=0.01 (95%CI:-0.01; 0.02)); compared to the SOC (ICI=-0.05 (95%CI: -0.07; -0.02). Between group comparison (*p*<0.001).

*Interpretation:* DAT interventions were cost-saving and reduced the inequitable distribution of patient costs compared to the SOC. This highlights the potential value of interventions that reduce health service visits in improving the equitable distribution of health services.

**Funding:** Unitaid (Grant Agreement Number: 2019-33-ASCENT).

**Research in context:** *Evidence before this study:* In November 2022, we searched PubMed and MedRxiv for English-language studies published between January 2000 and current, using the terms “tuberculosis” AND “cost” AND (“Digital Adherence Technologies” OR “DATS” OR “99DOTS” OR “Pillbox”). This search was repeated as part of a systematic review in April 2023 followed by an update in May 2024. Twenty-nine relevant studies have been identified, estimating the costs of DATS, though many did not assess the full economic costs of implementation. Only two studies included an assessment of patient-incurred costs, and none considering the equity distribution of costs or outcomes using an asset-based index.

*Added value of this study:* The ASCENT study provides robust evidence using a comprehensive economic evaluation framework, that DATs decreased the cost of tuberculosis treatment in Ethiopia for a cohort of adults with pulmonary tuberculosis. There was a 49-56% probability of DATs improving the cost-effectiveness of tuberculosis treatment and there was no significant difference in disability adjusted life years (DALYs) between study arms. The implementation of DATs did not change the distribution of costs or DALYs between people with tuberculosis (PWTB) of different household socio-economic position, however it did reduce the magnitude of patient costs among PWTB in the lower socio-economic position (SEP) quintile.

*Implications of all the available evidence:* While there is limited evidence of the effectiveness of Digital Adherence Technologies (DATs), this study is the first to show what impact the DATs may have on the costs of treatment, by reducing the number of healthcare visits leading to cost savings. There is further evidence that DATs may reduce the burden of patient costs on those who are least wealthy. We recommend that future investments in DATs for tuberculosis treatment support consider how healthcare providers integrate DATs for tuberculosis treatment support in the health facility workflow and how this translates to cost savings.

## Introduction

The 2018 World Health Assembly on Digital Health highlighted the potential of digital technologies to advance universal health coverage, while cautioning that contributions should be carefully assessed to ensure investments do not divert resources from alternative, more cost-effective non-digital approaches [1]. Tuberculosis, a notifiable disease of public health importance, requiring adherence to treatment schedules of 6 months or more, may benefit from innovation to support treatment adherence. In 2022, 7.5 million people globally started treatment for tuberculosis, and with an estimated 1.3 million deaths, it remains one of the deadliest infectious diseases [2]. In Ethiopia the tuberculosis incidence rate is 126 (85-176) cases per 100,000 population compared to the global average of 133. Disease risk factors include undernourishment, HIV, alcohol use disorders, diabetes, and smoking. Of individuals newly started on tuberculosis treatment in 2021, 87% successfully completed. Current treatment options for drug-susceptible tuberculosis (DS-TB) include a six-month course of treatment or a 6-20-month course for rifampicin-resistant tuberculosis (RR-TB). Intermittent dosing of tuberculosis medication as is observed with suboptimal dosing is possible due to the post-antibiotic effect of drugs included in first-line treatment. However, the treatment regimen will be less bacteriostatic when dosing do not coincide with the metabolic activity of semi-dormant persistent tubercle bacilli [3]. Consistent adherence is therefore recommended to reduce the risk of treatment failure, relapse and acquired drug resistance, especially among persons with initial pulmonary cavitation or HIV disease.

Historically, directly observed therapy short course (DOTS) was used to achieve high rates of treatment success, necessitating persons with TB (PWTB) attending the health facility daily to be observed when taking their medication. Digital solutions for adherence monitoring has made community-based, patient-centered treatment adherence support possible. A range of digital adherence technologies (DATs) have been assessed with limited evidence of improvements in end of treatment outcomes [4–6] and cost-effectiveness [7–10]. Interventions range from video-supported therapy and digital pillboxes to SMS-based interventions. The use of these interventions may be dependent on PWTB’s access to digital devices, and concerns have been raised about equity implications [11]. Since lower household socio-economic position (SEP) increases individuals’ risk of contracting tuberculosis disease [12], simultaneously increasing the risk for suboptimal treatment adherence and poor health outcomes, it is important to assess the equity implications of digital interventions. As part of the adherence support coalition to end tuberculosis (ASCENT) project, a pragmatic cluster randomised trial was conducted to assess the effectiveness and cost-effectiveness of digital pillboxes (pillbox) and medication labels (labels) with daily monitoring using a web-based platform to inform differentiated care, in reducing a composite unfavourable outcome, including tuberculosis recurrence in Ethiopia. The trial did not find evidence of a difference in the composite unfavourable outcomes between the labels (adjusted odd ratio [aOR]=1.14; 95%CI:0.83-1.61; *p*=0.62) or pillbox study arms (aOR=1.04; 95%CI:0.74-1.45; *p*=0.95) when compared against the standard of care (SOC) [13]. We conducted a trial-based distributional cost-effectiveness analysis to assess the equity impact, costs and cost-effectiveness of digital adherence technologies with differentiated care compared to the SOC in Ethiopia.

## Methods

### Study design and population

We conducted a trial-based distributional cost-effectiveness analysis using participant-level data, following a pre-specified health economics analysis plan [14]. Costs and outcomes were assessed from a societal perspective. ASCENT was a three-arm pragmatic cluster randomised trial of digital adherence technologies (DATs) for tuberculosis treatment support, followed by differentiated care compared to the standard of care (SOC) in Ethiopia. The unit of randomisation was health facilities, with 78 facilities (26 per study arm) in the Addis Ababa and Oromia regions of Ethiopia enrolled. Clusters were randomised (1:1:1) using stratified restricted randomisation, based on province and TB notifications to provide either SOC, medication labels (labels), or digital pillboxes (pillbox; EvriMed1000). 3,858 individuals ≥ 18 years of age, with pulmonary DS-TB were enrolled and followed up 12 months after treatment initiation to determine outcomes. Two trial endpoint measures were collected, (i) end-of-treatment outcome recorded in the facility TB treatment register, and (ii) for all participants with bacteriologically confirmed tuberculosis who had either a cured or completed end of treatment outcome, self-reported TB retreatment. For participants able to provide a sputum sample, culture was done six months after the end of treatment to measure disease recurrence. Data on patient events and trial endpoints were collected for all trial participants. Provider – and patient costs were collected from the same sample of 15 health facilities (5 per study arm), and 10 participants per facility (total of 150 observations).

### Ethics

The study received ethics approval from LSHTM Ethics Committee, UK (19120–1); WHO Ethical Review Committee, Switzerland (0003297); the Addis Ababa City administration health bureau public emergency and health research directorate institutional review board (A/A/H/B/947/227); and the Oromia Regional Health Bureau public emergency and health research directorate institutional review board (BEFO/HBTFH/1–16/10322), Ethiopia. Written informed consent were sought from individuals enrolled in the study.

### Interventions

Two DAT interventions, pillbox and labels, were introduced to health facilities randomised to the intervention arms (one DAT per intervention arm). Health facilities assigned to the SOC study arm continued routine practice. TB focal staff providing tuberculosis care in each of the health facilities, received training from trial staff in tuberculosis treatment adherence monitoring and the use of the DATs. Each dose taken by participants was logged on the platform either automatically when the participant opened the pillbox to take their medication, or when the participant texted a code on the dose label to the dedicated toll-free number. Participants receiving care in facilities randomised to labels, who did not have a phone were offered a pillbox. DATs were linked to an online platform for daily monitoring of participant engagement with the DAT. Automated SMS reminders were sent to all participants who did not open their pillbox or had not sent a text by 11:00 each day. Doses not recorded were considered missed and the TB focal person started a differentiated response based on data on the platform. The response included a phone call to the patient if 1-2 doses are missed; home visits by a community healthcare provider if there is evidence of persistent missed doses (>5 consecutive doses); and a switch to DOT was considered if >14 doses were missed.

### Assessment of household socio-economic position, cost and outcomes

Between May 2021 and August 2022, adults receiving tuberculosis treatment at participating health facilities were offered enrolment into the trial. Participants were followed up for 12 months until August 2023. Patient events were collected at enrolment, from treatment registers and at the 12-month follow-up [14]. Household socio-economic position (SEP) as a relative measure of poverty was estimated by calculating an asset index through principal component analysis of 27 multi-dimensional poverty indicators collected using a survey at enrolment (table S2). Using this index, participants were divided into quintiles of relative wealth. Costs and outcomes were compared between quintiles using concentration index-based indices [15]. Illness concentration curves were constructed by plotting the cumulative percentage of illhealth against the proportion of the population ranked by household SEP. The standard illness concentration index (ICI) derived from the curve was defined as twice the area between the concentration curve and the line of equality on the graph [16].

Cost categories included technology, network – and support costs, health service costs of treatment, and patient-incurred costs. Technology costs included the cost of the devices, accounting for cluster-level re-use rates for pillboxes. Network costs for labels was the costs of two-way SMS’ and monthly hosting costs of the web-based platform for both technologies. Support costs include the cost of training, support visits to health facilities, and 0.5 full-time equivalent of a technical officer to support the implementation and integration of the intervention. Provider costs were estimated at facility level for 13 health facilities. The cost of staff time was estimated using timesheets completed by tuberculosis focal staff (table S3). A separate log kept by trial staff was used to record the number of support visits, telephone calls and travel to health facilities. Time spent was valued using the average wage rate for health care workers in Ethiopia. The number of visits to health facilities were recorded prospectively for each participant, on a facility log and multiplied by the provider unit costs to calculate total costs. Costs valued before 2023 were inflated using the average annual consumer inflation rate in Ethiopia over the previous five years (22.2% per annum). For the cost of hospitalisation, diagnostic tests and RR-TB treatment, we used secondary data on unit costs multiplied by the number of events collected for each participant [17]. Data on patient-incurred costs such as transport and other out-of-pocket costs were collected by surveying 10 trial participants in each of the 15 health facilities (5 per study arm). Costs reported in Ethiopian Birr were converted to 2023 United States Dollars using the average annual exchange rate (1 EB = 0.018 USD).

We estimated participant-level disability-adjusted life years (DALYs) based on participants’ gender, age, and trial endpoint. DALYs are the product of years of life lost, and years lived disabled due to tuberculosis. Years of life lost due to premature mortality were based on age- and gender-stratified expected life expectancy for Ethiopia, if the participant died during treatment or prior to 12-month follow-up. Years lived disabled were calculated based on time spent on treatment (including recurrence or drug-resistant tuberculosis) multiplied by the global burden of disease disability weight for tuberculosis disease (Eq S6) [18].

### Statistical analyses

Cluster-level mean provider costs from 15 health facilities were singly imputed to the 78 trial health facilities by province (Addis/Oromia) and study arm. Patient costs, socio-economic position and DALYs at 12 months were jointly imputed for participants with missing values using multi-level multiple imputation by chained equations, generating 50 imputed datasets. For each of the imputed datasets, costs and DALYs were jointly modelled using bivariate Bayesian hierarchical models and log-normal distributions with random effects to allow for cluster-level variances. Data were fitted using Markov Chain Monte Carlo sampling to obtain posterior simulations. The results of the imputed models were pooled by combining the posterior draws for all 50 imputed models. Total costs and DALYs in each study arm were estimated by summing participant-level costs and DALYs. The mean differences in costs and DALYs were calculated group-wise by subtracting the arithmetic mean cost in each of the intervention groups from those in the SOC arm. The incremental cost-effectiveness ratio (ICER) was calculated by the differences in costs divided by the difference in DALYs averted between the intervention arms and the SOC study arm (Eq S7). The distributional incremental cost-effectiveness ratio (dICER) was calculated as the ratio of net health benefit divided by the difference in the illness concentration indices (ICI) between the intervention and SOC study arms (Eq S11). ICERs were compared against a cost-effectiveness threshold for Ethiopia of $100 per DALY averted [19].

Prespecified sensitivity analyses were used to assess how changes in the analysis may affect the results of our study; including (i) changing the number of healthcare visits in the SOC to the number of visits observed under guidelines circumstances; (ii) increasing the cost of the intervention; (iii) only including the costs and outcomes incurred during the treatment of trial participants until the end of the trial; (iv) valuing participant time using the average wage rate of government employees in Ethiopia; and (v) varying the discount rate used for costs and effects from 0% to 10%. Variance in the posterior draws of the model compared against a range of thresholds were used to quantify the uncertainty in our estimates in a cost-effectiveness acceptability frontier for the probabilistic sensitivity analysis. Data were analysed using Stata 18 and R statistical software. The study is reported using the Consolidated Health Economic Evaluation Reporting Standards (CHEERS) 2022 checklist.

### Governance

The study was funded by UNITAID (2019-33-ASCENT) through the Adherence Support Coalition to End TB (ASCENT) project. The funder had no role in study design, data collection, data analysis or interpretation or the writing of the manuscript. The trial had independent oversight from a Technical Advisory Group (TAG) with representatives from all five ASCENT countries, as well as a Community Advisory Group (CAB) including representatives from people affected by tuberculosis in Ethiopia. The Ethiopian National TB Programme was involved in the design of the economic evaluation and received regular updates. The trial is registered with Pan African Clinical Trial Registry (PACTR) PACTR202008776694999, registered on 11 August 2020 at https://pactr.samrc.ac.za/TrialDisplay.aspx?TrialID=12241.

## Results

Between May 2021 and August 2022, 3885 adults (≥18 years) starting DS-TB treatment were enrolled, with follow-up visits completed in August 2023 (figure S1). Participants were excluded if their diagnosis changed to “not TB” or if they were diagnosed with RR-TB treatment less than 28 days before treatment start or were re-enrolled leaving 1295 participants in the standard of care (SOC), 1305 in the Labels -, and 1258 in the Pillbox arms of the study in the intention-to-treat population. Participants were analysed in the based on DAT received, leaving 1295 in SOC, 1152 in labels and 1411 in pillbox (table S1). Baseline characteristics of the clusters and trial participants are shown in Table 1.

**Table 1.** Baseline characteristics of the enrolled population by study arm participants received.

|  | Standard of<br>care | Labels | Pillbox |
| --- | --- | --- | --- |
| **Characteristics of the clusters/ health facilities** |  |  |  |
| Number of clusters | 26 | 26 | 26 |
| Region |  |  |  |
| Addis Ababa | 12 (46%) | 12 (46%) | 12 (46%) |
| Oromia | 14 (54%) | 14 (54%) | 14 (54%) |
| Area |  |  |  |
| Urban | 24 (92%) | 24 (92%) | 25 (96%) |
| Rural | 2 (8%) | 2 (8%) | 1 (4%) |
| Median number of individuals per cluster<br>(IQR) | 48 (41, 59) | 50 (36, 65) | 45 (39, 50) |
| **Baseline characteristics of the enrolled population** |  |  |  |
| Number of trial participants | 1 295 | 1 152 | 1 411 |
| Mean age in years (SD) | 35.5 (14.7) | 32.4 (13.5) | 34.3 (14.1) |
| Female | 496 (38%) | 487 (42%) | 584 (41%) |
| Bacteriologically confirmed tuberculosis | 806 (62%) | 756 (66%) | 936 (66%) |
| Previous tuberculosis treatment | 112 (9%) | 63 (5%) | 81 (6%) |
| HIV status |  |  |  |
| Negative | 1 134 (88%) | 1 012 (88%) | 1 218 (86%) |
| Positive, not on ART | 2 (0%) | 4 (0%) | 9 (1%) |
| Positive, on ART | 157 (12%) | 133 (12%) | 182 (13%) |
| Unknown | 2 (0%) | 3 (0%) | 2 (0%) |
| Educational level |  |  |  |
| None (Illiterate) | 198 (15%) | 114 (10%) | 220 (16%) |
| Less than primary | 73 (6%) | 65 (6%) | 119 (8%) |
| Primary | 635 (49%) | 571 (50%) | 647 (46%) |
| Secondary or Higher | 376 (29%) | 394 (34%) | 419 (30%) |
| Unknown | 13 (1%) | 8 (1%) | 6 (0%) |
| Mean number of people per room of the<br>house (SD) | 2.6 (1.7) | 2.6 (1.6) | 2.5 (1.5) |
| Relationship status and living arrangements<br>as a proxy for social capital |  |  |  |
| Single | 309 (24%) | 313 (27%) | 218 (15%) |
| Single (independent) | 193 (15%) | 211 (18%) | 321 (23%) |
| Married or cohabitating | 621 (48%) | 528 (46%) | 146 (10%) |
| Separated or widowed | 158 (12%) | 93 (8%) | 52 (4%) |
| Unknown | 14 (1%) | 7 (1%) | 6 (0%) |
| Frequency of income received |  |  |  |
| No income | 277 (21%) | 266 (23%) | 324 (23%) |
| Irregularly | 198 (15%) | 127 (11%) | 286 (20%) |
| Seasonally | 243 (19%) | 245 (21%) | 291 (21%) |
| Monthly | 563 (43%) | 507 (44%) | 504 (36%) |
| Unknown | 14 (1%) | 7 (1%) | 6 (0%) |
| Household socio-economic position |  |  |  |
| Poorest | 253 (20%) | 211 (18%) | 263 (19%) |
| Less poor | 258 (20%) | 187 (16%) | 280 (20%) |
| Middle | 227 (18%) | 209 (18%) | 289 (20%) |
| More wealthy | 239 (18%) | 238 (21%) | 249 (18%) |
| Wealthiest | 211 (16%) | 260 (23%) | 257 (18%) |
| Unknown | 107 (8%) | 47 (4%) | 73 (5%) |

All data shown are n(%) or mean (SD) or n/N(%). IQR=interquartile range. ART=antiretroviral therapy. SD=standard deviation. Each health facility is one cluster.

Baseline characteristics were similar between study arms. 62% (806/1295) of participants in the SOC, compared to 66% (756/1152) in the labels and 66% (936/1411) in the pillbox arms had bacteriologically confirmed tuberculosis. Slightly more participants in the SOC arm had a history of previous tuberculosis disease at 9% (112/1295) compared to 5% (63/1152) in the labels and 6% (81/1411) in the pillbox arms. Considering the variables describing household socio-economic position (SEP), similar proportions of participants in the SOC 21% (277/1295) compared to the labels 23% (266/1152) and pillbox 23% (324/1411) arms of the study reported no individual income. Participants in the SOC arm were less wealthy (20% classified as lowest wealth quintile) compared to the labels and pillbox arms (18% and 19% respectively). Participants in the labels study arm were wealthier (43% (498/1152) in the two wealthiest quintiles) than those in the SOC (35% (450/1295)) and pillbox (36% (506/1411)) study arms. The number of health facility visits per treatment episode compared between study arms are shown in table S4. Compared to the SOC, there was a reduction in the mean number of visits to health facilities in the pillbox and labels study arms throughout the treatment period; labels compared to SOC by -20.2 (95%CI: -26.3; -14.1; *p*<0.001) and pillbox compared to SOC -20.4 (95%CI: -26.5;-14.3; *p*<0.001) (table S4). Cohort level patient events during treatment compared between study arms are summarised in table S5. There were more home visits in the labels (77), and pillbox (58) compared to the SOC (5) study arms. Similarly nights in hospital was greater in the labels (648) and pillbox arms (310) compared to the SOC (135). This was partly explained by one healthfacility in the labels arm that admitted more people to hospital, and may also be as a result of more intensive follow-up because of monitoring using the DATs. A total of 22 participants required a second course of tuberculosis treatment in the SOC, compared to 24 in the labels and 31 in the pillbox study arms. With 4 participants in the SOC, 1 in labels and 2 in the pillbox study arms starting rifampicin resistant tuberculosis treatment. The economic costs of treatment are summarised in Table 2.

**Table 2.** Total and per-patient costs (2023 USD) incurred for trial participants, by cost categories and study arm.

|  | Standard of Care (n=1295) |  | Labels (n=1152) |  | Pillbox (n=1411) |  |
| --- | --- | --- | --- | --- | --- | --- |
|  | Total costs [95%CI] | Cost/patient [95%CI] | Total costs [95%CI] | Cost/patient [95%CI] | Total costs [95%CI] | Cost/patient [95%CI] |
| Technology | N/A | N/A | \$806 [fixed] | \$0.70 [fixed] | \$11,006 [10,752; 11,161] | \$7.80 [7.62; 7.91] |
| Network costs | N/A | N/A | \$12,534 [fixed] | \$10.88 [fixed] | \$9,665 [fixed] | \$6.85 [fixed] |
| Support and training | \$1,334 [1,308; 1,373] | \$1.03 [1.01; 1.06] | \$1,140 [1,117; 1,175] | \$0.99 [0.97; 1.02] | \$1,270 [1,242; 1,298] | \$0.90 [0.88; 0.92] |
| **Sub-total intervention costs** | **\$1,334 [1,308; 1,373]** | **\$1 [1; 1]** | **\$14,481 [14,135; 14,826]** | **\$13 [12; 13]** | **\$21,941 [21,419; 22,364]** | **\$16 [15; 16]** |
| Health facility visits | \$472,675 [448,070; 498,575] | \$365 [346; 385] | \$78,336 [76,032; 79,488] | \$68 [66; 69] | \$88,893 [87,482; 91,715] | \$63 [62; 65] |
| Drug sensitive tuberculosis treatment | \$45,325 [44,030; 45,325] | \$35 [34; 35] | \$40,320 [39,168; 40,320] | \$35 [34; 35] | \$45,325 [47,974; 49,385] | \$35 [34; 35] |
| Smear microscopy tests | \$75,110 [75,110; 76,405] | \$58 [58; 59] | \$66,816 [66,816; 67,968] | \$58 [58; 59] | \$81,838 [81,838; 83,249] | \$58 [58; 59] |
| Hospitalisation | \$1,036 [376; 1,645] | \$0.80 [0.29; 1.27] | \$4,712 [2,684; 6,751] | \$4.09 [2.33; 5.86] | \$2,300 [1,242; 3,344] | \$1.63 [0.88; 2.37] |
| Treatment for recurrence (drug-sensitive) or multi-drug resistance tuberculosis | \$41,440 [6,475; 76,405] | \$32 [5; 59] | \$19,584 [1,152; 40,320] | \$17 [1; 35] | \$28,220 [1,411; 55,029] | \$20 [1; 39] |
| **Sub-total provider costs.** | **\$635,586 [574,061; 698,355]** | **\$491 [443; 539]** | **\$209,768 [185,852; 234,847]** | **\$182 [177; 204]** | **\$246,576 [219,947; 282,722]** | **\$178 [156; 200]** |
| Patient incurred costs<sup>1</sup> | \$10,360 [10,360; 10,360] | \$8 [8; 8] | \$10,368 [9,216; 10,368] | \$9 [8; 9] | \$9,877 [8,466; 9,877] | \$7 [6; 7] |
| Productivity costs<sup>2</sup> | \$10,360 [10,360; 10,360] | \$8 [8; 8] | \$6,475 [6,475; 6,475] | \$5 [5; 5] | \$7,055 [7,055; 8,466] | \$5 [5; 6] |
| **Sub-total patient-incurred costs** | **\$20,720 [20,720; 20,720]** | **\$16 [15; 16]** | **\$16,843 [15,691; 16,843]** | **\$14 [13; 14]** | **\$16,932 [15,521; 18,343]** | **\$12 [11; 13]** |
| **Total societal costs** | **\$657,640 [596,089; 720,447]** | **\$507 [458; 555]** | **\$241,091 [215,678; 266,516]** | **\$196 [190; 218]** | **\$285,449 [256,887; 323,429]** | **\$206 [167; 213]** |

All costs are shown as the mean costs in 2023 USD. <sup>1</sup>Based on single imputation. <sup>2</sup>Productivity costs are based on the value of individuals' time spent travelling to health facilities or waiting at the facility. 95%CI=95% confidence interval. Technology and network costs are fixed costs per patient as primarily purchased from third party providers at a fixed rate of cost per patient, except in the technology costs for pillbox where there is variability because of differences in cluster-level device re-use rates.

Intervention costs were $13 (95%CI: 12; 13) per patient in the Labels compared to $16 (95%CI: 15; 16) in the Pillbox arm, with an empirically derived pillbox reuse rate of 2.16 (95%CI: 1.58; 2.85) included in the calculation. Per patient provider costs were much higher in the SOC $491 (95%CI: 443; 539) compared to the Labels $182 (95%CI: 177; 204) and pillbox study arms $178 (95%CI: 156; 200), driven by the larger number of health care visits in the SOC. The cost of medication, diagnostic tests, hospitalisation, and further treatment were similar between study arms. Patient-incurred costs are similar between study arms at $16 (95%CI 15; 16) per person in the SOC, $14 (95%CI: 13; 14) and $12 (95%CI: 11; 13) in the labels and pillbox arms respectively despite the large difference in the number of health facility visits in the intervention arms. The distributions of DALYs, health facility visits, patient – and provider costs by household socio-economic position (SEP) are shown in Figure 1.

**Figure 1.**
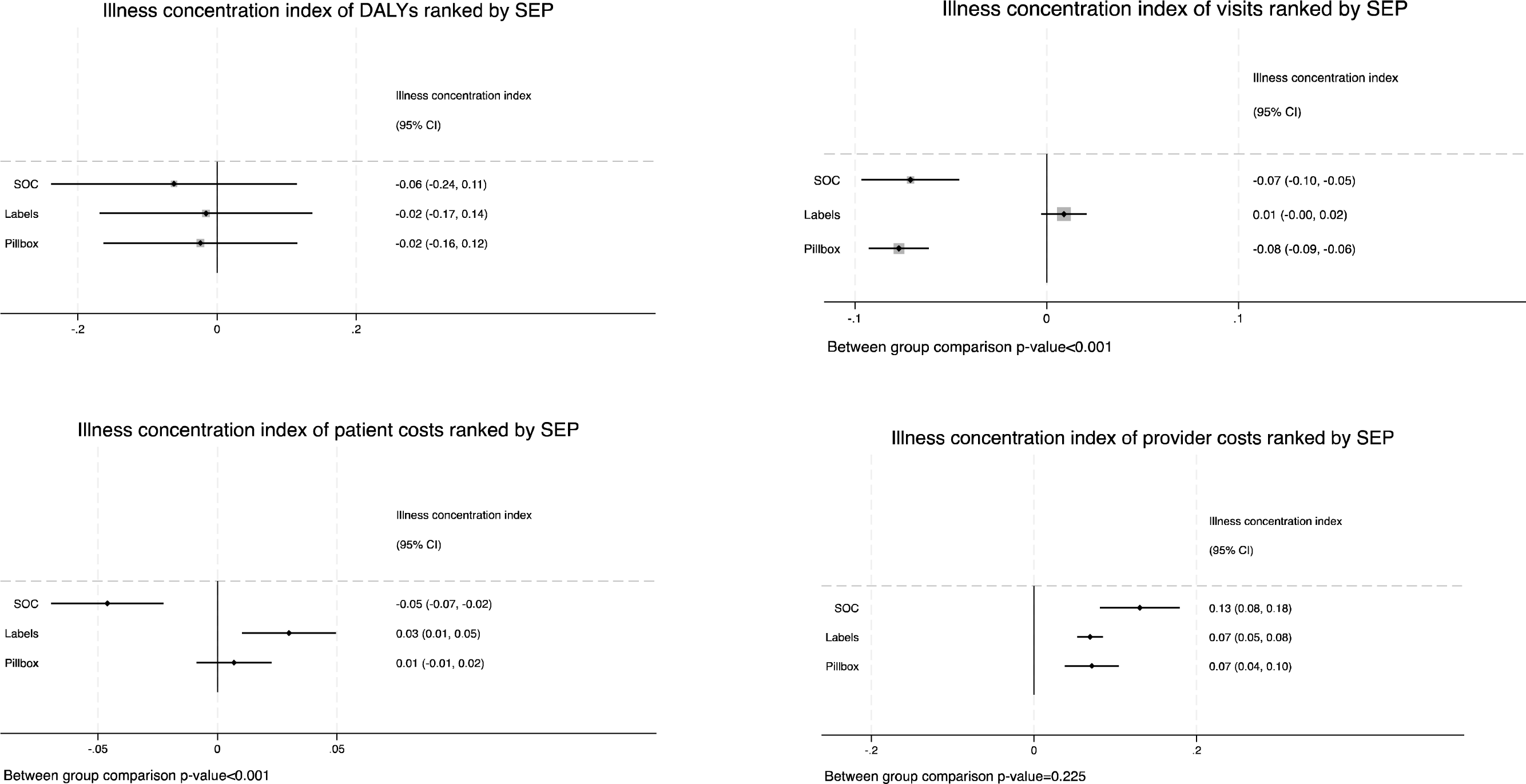
illness concentration indices for (A) Disability Adjusted Life Years (DALYs); (B) Number of health facility viists; (C) Patient costs in 2023 USD; and (D) Provider costs in 2023 USD. The concentration index (ICI) assesses if there is a within study arm difference in the outcome by ranked by household socio-economic position (SEP). Clustering was taken into account in estimating 95% confidence intervals (95%CI). A negative concentration index means that the outcome, here a negative consequence, is concentrated among less wealthy households. Conversely, a positive index value means that the outcome is concentrated among relatively wealthier households. We compared the difference in the distribution in outcome between SEP groups, where the p-value represents the likelihood of any observed difference between the groups being by chance.

The equity impact of the interventions in four domains, namely DALYs, health facility visits, patient-incurred costs and provider costs are shown in figure 1, figure s2 and table S6. The illness concentration indices (ICIs) for DALYs were -0.06 (95%CI: -0.24; 0.11) in SOC; -0.02 (95%CI: -0.17; 0.14) in labels and -0.02 (95%CI: -0.16; 0.12) in pillbox study arms, with no evidence of differences in the distribution of DALYs within or between study arms (*p*=0.920). The number of health facility visits were concentrated among the comparatively poor in the SOC -0.07 (95%CI: -0.10; -0.05) and pillbox - 0.08 (95%CI: -0.09; -0.06) arms but not in the labels arm 0.01 (95%CI: -0.00; 0.02). Patient costs were concentrated among those relatively poor in the SOC arm -0.05 (95%CI: -0.07; -0.02), while concentrated among the wealthy in the labels arm 0.03 (95%CI: 0.01; 0.05) and pillbox 0.01 (95%CI: - 0.01; 0.02) with evidence of differences between study arms (*p*<0.001). Provider costs were concentrated among the relatively wealthy, suggesting that the wealthy most benefits from public health expenditure, in the SOC 0.13 (95%CI: 0.08; 0.18); labels 0.07 (95%CI: 0.05; 0.08) and pillbox 0.07 (95%CI: 0.04; 0.10) arms, though there was insufficient evidence (*p*=0.225) of a difference between study arms. The cost-effectiveness and distributional cost-effectiveness of the interventions compared to the SOC are summarised in Table 3.

**Table 3.** Incremental cost-effectiveness from the provider – and societal perspectives.

|  | Simple imputation of the means |  | Multilevel multiple imputation |  | Group-level summary metrics |  |
| --- | --- | --- | --- | --- | --- | --- |
|  | Incremental costs per patient [95% CI] | DALYs averted [95% CI] | Incremental costs per patient [95% CI] | DALYs averted [95% CI] | Incremental cost-effectiveness ratio [95% CI] | Incremental distributional cost-effectiveness ratio [95% CI] |
|  | Mean difference [95% CI] | Mean difference [95% CI] | Mean difference [95% CI] | Mean difference [95% CI] |  |  |
| <u>Provider perspective</u> |  |  |  |  |  |  |
| Labels versus SOC | -309 [-266; -335] | -0.46 [-2.07; 1.17] | 7 [-191; 189] | 0.04 [-1.93; 1.93] | 672 [14; 1420] | 18.53 [4.16; 31.85] |
| Pillbox versus SOC | -313 [-287; -339] | -0.32 [-1.61; 0.97] | -1 [-190; 183] | -0.01 [-1.93; 1.93] | 978 [-54; 1626] | 19.80 [8.88; 30.73] |
| <u>Societal perspective</u> |  |  |  |  |  |  |
| Labels versus SOC | -311 [-268; -337] | -0.46 [-2.07; 1.17] | 7 [-191; 189] | 0.04 [-1.93; 1.93] | 676 [17; 1435] | 18.68 [4.30; 31.99] |
| Pillbox versus SOC | -301 [-291; -342] | -0.32 [-1.61; 0.97] | -1 [-190; 183] | -0.01 [-1.93; 1.93] | 941 [-53; 1652] | 18.96 [9.16; 30.94] |

\*Costs reported based on multiple multi-level imputation. In a separate analysis, we estimated costs using single imputation where the means of patient costs were used from the subsample – the findings of this analysis are summarised in the appendix along with a detailed report of the costs estimated. The current estimates suggest that the cost-effectiveness threshold (CET) for Ethiopia is between \$10 and \$255 (we used an average of \$93) per DALY averted. For the comparison, we use \$100 as an estimate within the range. LTFU: lost to follow-up. <sup>f</sup>Mean age at death was 48.6 (SD: 18.6) years in the Standard of Care arm; 43.9 (SD: 17.8) years in the Labels - and 41.2 (SD: 14.7) in the pillbox arm. DALYs: Disability adjusted life years. SD: standard deviation. 95%CI: 95% Confidence Interval. The distributional cost-effectiveness ratio was estimated, with the efficiency impact being the net monetary benefit calculated as the change in effectiveness multiplied by the CET minus the change in costs (EqS8); and the equity impact being the difference in health inequality measured as the difference in the illness concentration indices between study arms. The distributional cost-effectiveness ratio is then a ratio of incremental efficiency/ equity impact (EqS12).

The difference in societal costs, using multiple imputation, was a reduction by $299 per patient in the Labels and $298 in the Pillbox arms when compared against the SOC. The societal cost per DALY averted was $676 (95%CI 17; 1435) in the Labels and $941 (95%CI -53; 1652) in the Pillbox study arms.

**Figure 2.**
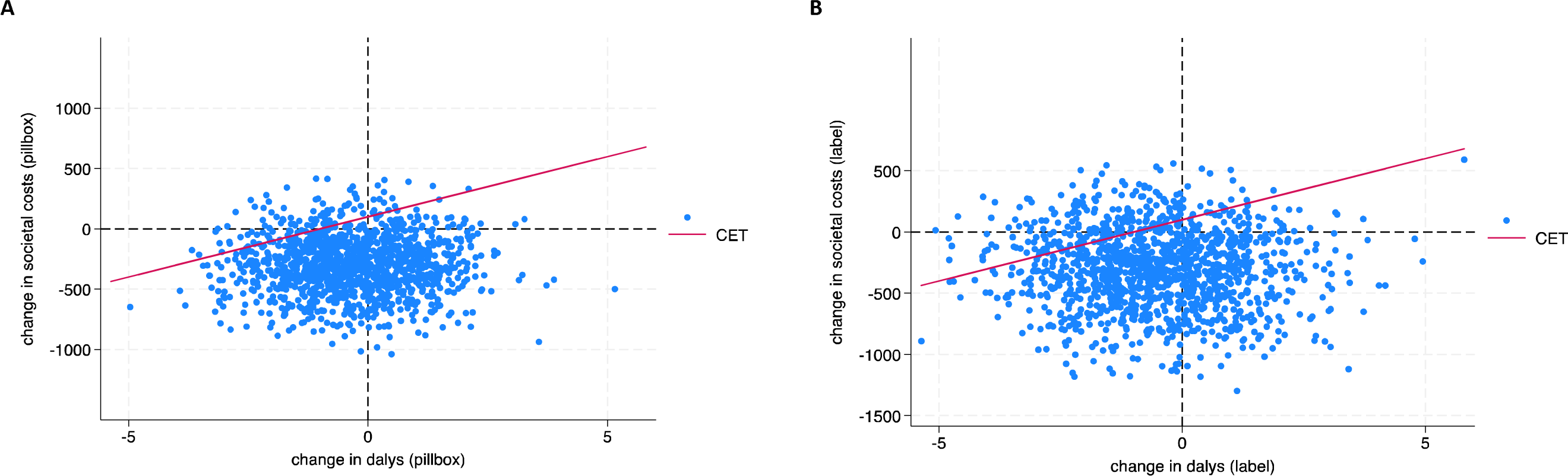
Cost-effectiveness planes. Scatter plot of the estimated joint density of incremental costs and incremental DALYs averted, showing uncertainty in the ICER estimate obtained from the analysis models. The dots represent the findings of a probabilistic sensitivity analysis.

When plotted on cost-effectiveness planes, we find that the ICERs are crossing the axes but slightly concentrated in the bottom left hand quadrant suggesting that DATs are less costly and less effective than the standard of care. ICERs in this quadrant are interpreted as cost-effective if they fall below the cost-effectiveness threshold. Using the outcomes of the probabilistic sensitivity analysis, plotted on the cost-effectiveness acceptability frontier, the probability that each of the interventions would be cost-effective compared against a range of cost-effectiveness thresholds (CETs) are shown in figure S3. Using a range of US$5 to US$900 per DALY averted for the CETs, we find that the interventions have a 56% probability (labels) and 49% probability (pillbox) of being cost-effective compared to SOC at a CET of US$100. The results of the distributional cost-effectiveness analysis estimates the distributional cost-effectiveness ratio at 18.68 for labels and 18.96 for pillbox. We compare this finding against a relative health inequality aversion parameter of 5.66. In the absence of empirical research on the level of inequality aversion by policymakers in Ethiopia, we used an estimate previously derived for decisions in Ethiopia [20]. As the value of the inequality aversion parameter increases, it suggests that there is a greater willingness to achieve fewer health improvements to reduce inequality in health or other outcomes. Our distributional cost-effectiveness ratio estimated (18.68 and 18.96) exceeds the parameter used for previous decisions in Ethiopia for vaccines (5.66) suggesting that policy makers would need to be more averse to inequality than in previous decisions, to recommend the implementation of DATs based on health equity.

In the sensitivity analyses (table S7), we found that more visits in the SOC increased potential cost-savings from implementing the interventions. If we assumed an increased cost of the pillbox, for example in setting where reuse rate is low, the cost savings compared to the SOC reduced by 16%. Removing the costs and DALYs associated with recurrence reduces cost savings and has minimal effect on the DALYs averted. Valuing participants using an average wage rate increases the cost savings to patients. Main study findings, however, remained similar under sensitivity analysis.

## Discussion

In health facilities implementing DATs, trial participants made 20 fewer visits per treatment episode compared to the standard of care (SOC), substantially reducing the cost to providers. We didn’t find evidence of differences in poor health outcomes (DALYs) between study arms or their distribution between socio-economic position (SEP) groups. However, there was a reduction in patient-incurred costs among the least wealthy households in the intervention compared to SOC study arms. When assessing uncertainty, the 95% confidence intervals of the incremental cost-effectiveness ratios (ICERs) were wide, suggesting substantial uncertainty that DATs would be cost-effective when compared to the SOC. We estimated that the labels and pillbox arms have a 49-56% probability of improving the cost-effectiveness of tuberculosis treatment compared to SOC at a cost-effectiveness threshold of $100 per DALY averted. Our study follows the work of other cluster randomised trials evaluating the effectiveness and costs of Digital Adherence Technologies (DATs) in China [5,10], South Africa [21], Uganda [7] and Ethiopia [22], by including a detailed analysis of human resources, patient costs and equity in addition to costs and cost-effectiveness.

Differences in provider costs between the SOC and intervention arms of the study were primarily driven by the number of healthfacility visits per treatment episode. In a separate analysis, we showed that health care workers spent more time with PWTB per visit in the intervention arms than when they were attending more frequently in the SOC arm [23]. This increase in time spent remained lower than the cumulative time of multiple visits in the SOC. Our evaluation of cost of treatment included above service-level costs. These costs were highly variable, particularly between urban and rural health facilities where fewer people with tuberculosis (PWTB) were accessing services. Given that we had overhead costs from only 12 clusters, single imputation of provider costs has likely resulted in larger variability than if we had collected data for all 78 clusters. Provider costs reported in this study were broadly similar to the costs estimated by Terefe et al’s comprehensive assessment of the cost of tuberculosis care in Ethiopia, though we used a micro-costing approach [17]. Our total cost estimate, includes costs incurred by all participants who received the intervention irrespective of whether they completed treatment (ITT population). Provider costs incurred through diagnostic tests, hospitalisation and subsequent treatment episodes were similar between study arms, and differences in treatment costs were predominantly driven by the reduction in the number of health facility visits.

Patient costs were similar in the intervention - compared to the SOC arms of the study, despite large differences in facility visits. This is explained by lower transport costs per visit reported in the SOC where there are more frequent visits compared to the intervention study arms. Furthermore, we used the human capital approach for valuing time spent seeking care, whereby individual self-reported income is used to value people’s time. This likely does not fully captured the full cost of time spent. Approximately 35% of participants reported no- or irregular income with consequently no cost allocated to their time spent. Furthermore, we collected self-reported income data at enrolment into the trial when people were on treatment and already had suffered disease-associated income loss. A recent meta-analysis of patient costs associated with tuberculosis in Ethiopia, found that pre-diagnosis costs were more than double post-diagnosis costs US$97.6 (56.4-184.3) versus US$45.1 (−119.1-209.37)) [28]. Differences in time spent between study arms due to a reduction in number of visits, would therefore only have a marginal effect on productivity costs. We quantified this potential bias by conducting a sensitivity analysis estimating the productivity costs for each of the study arms by valuing all participants’ time equally.

Our study is the first to empirically demonstrate that DATs could be cost saving by reducing the number of health facility visits. This observed difference is dependent on the frequency of health facility visits in the SOC. At the time of our study, PWTB were attending health facilities on average 30 times per treatment episode, our findings may therefore not hold in settings where less frequent facility visits are already routine practice. Furthermore, the observed difference between study arms would have been more pronounced prior to COVID-19, when visits to health facilities were more frequent [25]. Other studies evaluating the economic costs of DATs found them to be more costly and effect-neutral [7,8,10]. In Uganda, the provider cost of 99DOTS was estimated to be $59 (range: $50-$70) per participant successfully treated [7]. Similarly, in Morocco, the cost-effectiveness of a digital pillbox from a provider perspective was found to be $434 per DALY averted among MDR-TB patients. This was a model-based analysis where the number (and cost) of health facility visits were assumed to be the same between study arms [8]. A study in China, considering the societal perspective, found digital pillbox to be more costly and effect neutral with an ICER of $3,668.59 per DALY averted. Data for assessing number of health facility visits was limited (n=120) and did not fully assess differences in practice between studyarms [10].

Strengths of our study include it being a trial-based cost-effectiveness analysis with detailed data collection of patient events, healthfacility visits and rigourous assessment of trial endpoints embedded within a randomised study where the SOC reflects routine practice in Ethiopia. We furthermore went beyond the traditional cost-effectiveness framework to analyse the equity impact of the interventions. Study limitations include, the trial follow-up period was not long enough to capture all health and economic consequences of the interventions – particularly tuberculosis transmission. This question was addressed in a transmission modelling study following this study [26]. The number of health facility visits were recorded prospectively, and patient event data (including hospitalisations) collected at the 12 month follow-up. Self-reported patient events would not be available for patients who died or became LTFU during treatment episode and we underestimate the cost associated with care-seeking prior to death or LTFU. Given the representativeness of the sample, imputed costs would undervalue true costs. Our DALY estimates are likely to be conservative, because we only include DALYs for the current and identified future treatment episodes and not for any longer-term sequalae of tuberculosis disease [27]. Our estimates are similarly likely to underestimate the full costs and economic losses incurred because our analytical time horizon, restricted to the current tuberculosis event, includes costs incurred once participants started treatment and not during the diagnostic phase. We could only assess the equity distribution impact of the intervention among those already accessing care, and expecting to live in the health facility’s catchment area for the duration of treatment. We may therefore be underestimating the value of the intervention among more marginalised groups. Nevertheless, we expect the findings to be representative of adults who are accessing care at public health clinics in Ethiopia.

We used relative SEP based on multi-dimensional poverty assessment, to incporate equity in the decision framework [29]. Alternative approaches are based on estimates of individual income to assess inequality [30]. Measures of income-based inequality would correctly ascertain immediate ability to respond to a health shock, but would fail to account for resource-sharing within families, saving ability, and non-monetary assets that are important in assessing poverty in rural areas. Multi-dimensional poverty metrics include locally-relevant assets such as land, livestock, social capital and education that individuals may draw on in the event of a health shock. Our evaluation as part of the trial, was not powered to compare outcomes between SEP groups within study arms, but sufficient to evaluate how the concentration of the outcome between those relatively wealthy and relatively poor varies between study arms.

In conclusion, we found the implementation of DATs to be effect-neutral but cost-saving due to a marked reduction in healthfacility visits. These savings would only be fully realised if healthcare providers have other productive tasks to fill their time previously spent on DOTS, or in contexts where routine care is not daily facility-based DOTS. We found limited evidence of differences in DALYs averted between study arms and estimate a 49-56% probability that DATs will improve the cost-effectiveness of tuberculosis treatment at a CET of $100 per DALY averted. The implementation of DATs reduced the inequitable burden of patient-incurred costs.

## Authors’ contributions

NF, KLF, AWT and CFM designed this study. The trial was designed by KLF, KvK, JvR and DJ. All authors have contributed to the study design. KLF provided statistical expertise in clinical trial design and analysis. NF conducted the analysis and provided health economics expertise in design and analysis. AWT, MB, MS, AB, TA, NM, KvK and JvR implemented the study. All authors contributed to the interpretation of the findings and revision of the final manuscript. The authors read and approved the final manuscript.

## Acknowledgements

The authors acknowledge the contributions of the participants who enrolled in the study, the Ethiopian tuberculosis focal persons implementing the digital adherence technologies and the National Tuberculosis Program at the Ministry of Health Ethiopia without whom this study would not have been possible. We acknowledge the oversight and inputs of the ASCENT Technical Advisory Group.

## Data availability

Data analysed in this study are part of the ASCENT Consortium Trials data. A minimal dataset required to reproduce the findings and the survey materials is available from https://doi.org/10.17037/DATA.00003721 and analysis code from https://github.com/nicolacfoster. The minimal dataset is available for non-commercial use upon request, after signing a data sharing agreement, to studies approved by an ethics committee. Any publications arising from the shared data must acknowledge the investigators who collected the data, the institutions involved, and the funding sources by citing the data record and including a standard acknowledgement statement provided.

